# Exploring heterogeneity in mosquito exposure and attraction and its implications for malaria transmission

**DOI:** 10.1101/2025.09.04.25335091

**Authors:** Lars Kamber, Aurélien Cavelan, Melissa A Penny, Nakul Chitnis, Emma L Fairbanks

## Abstract

**Background:** Malaria transmission exhibits significant heterogeneity within communities, with small proportions of individuals experiencing disproportionate mosquito exposure.

**Methods:** This study addresses critical knowledge gaps in characterising and modelling this heterogeneity. Parameterising Bayesian hierarchical models to field data from Burkina Faso, we compared gamma and lognormal distributions for describing heterogeneity in mosquito biting rates. We then implemented this heterogeneity in an individual-based stochastic modelling platform, OpenMalaria, to assess its impact on transmission dynamics.

**Findings:** The gamma distribution was found to better describe the field data, compared to the lognormal distribution. When heterogeneity is introduced to malaria transmission simulations, both prevalence and incidence levels generally decrease across all age groups. Additionally, heterogeneity shifts disease burden towards younger age cohorts and alters the fundamental relationships between entomological inoculation rate and prevalence/incidence. The integration of appropriate heterogeneity distributions into transmission models substantially improved their ability to reproduce field-observed age-incidence curves.

**Interpretation:** Our findings highlight the importance of accounting for heterogeneous exposure when modelling malaria transmission, particularly in low-transmission and elimination settings where heterogeneity may be more pronounced.

## 1 Introduction

Malaria prevalence has been observed to vary not only between neighbouring villages, but also within villages [1, 2]. Like many diseases, a small proportion of the population often drives malaria transmission within communities. These ‘superspreaders’ are responsible for a significant portion of the transmission. A study analysing data from 90 African communities revealed that 20% of the population accounts for 80% of infections [3, 4]. This heterogeneity arises from multiple factors, including proximity to breeding sites, intervention usage, housing quality, pregnancy status, existing malaria infections, genetic factors and individual human behaviour [5–10]. However, the specific patterns and distributions of exposure heterogeneity at the household level remain poorly characterised. Understanding these patterns is crucial for accurately modelling transmission dynamics and evaluating control measures [11].

Many studies have observed heterogeneity in mosquito biting rates [12], however, there is a lack of comprehensive quantitative analysis to determine the implications of such heterogeneity. The expected number of infectious mosquito bites a human host receives is often referred to as the entomological inoculation rate (EIR). Data on the EIR is often collected to monitor the effectiveness on interventions, disease burden and vector species compositions [13, 14]. Previous research has identified age as a key factor in EIR heterogeneity [15, 16]. This is likely influenced in part by body surface area [17], which has been shown to be correlated with biting rates for other vectors and host species [18–20].

Many mathematical models for describing disease transmission dynamics assume homogeneous mixing patterns, which means that all individuals in a population have an equal probability of coming into contact with one another, regardless of their age, location, social connections or behavioural patterns. These assumptions often extend into vector-borne disease models, with each individual equally likely to be bitten by each vector. However, real-world disease transmission patterns exhibit significant heterogeneity, where contact rates and vector biting preferences vary based on factors such as age-structured social mixing, spatial clustering of populations, and vector feeding behaviours [21, 22]. Incorporating such heterogeneity into mathematical models can enhance the precision of predictions and policy recommendations.

While the importance of heterogeneous exposure in vector-borne disease transmission was recognised early on, the theoretical implications were first rigorously explored by Dye and Hasibeder [23]. Their work demonstrated that when mosquitoes concentrate on certain hosts, both the basic reproductive rate and vectorial capacity may be greater than their values under homogeneous mixing assumptions. Ross and Smith [24] demonstrated that transmission heterogeneity significantly influences all malaria epidemiological outcomes in simulation models. They found different patterns of heterogenity produce distrinct age-prevalence and incidence curves. These theoretical insights highlighted that ignoring heterogeneity in transmission models could lead to underestimating both disease persistence and the control efforts required for elimination [23].

Heterogeneity in biting rates between ages groups is often accounted for in simulations of OpenMalaria, an individual-based stochastic model describing malaria transmission dynamics, reducing the EIR of younger children [25]. However, it is also possible to simulate OpenMalaria assuming an additional heterogeneity in biting rates, where some individuals are more likely to be bitten, independent of age. Here, an OpenMalaria user can add this additional heterogeneity according to an lognormal distribution, specifying a mean and variance.

Our study builds on this foundation by addressing knowledge gaps of heterogeneity in malaria transmission, exploring possible distributions of this heterogeneity and its implications for malaria transmission in different settings. We compare the gamma and lognormal distributions to describe the heterogeneity between the rates at which individuals are bitten. Firstly, we fit previously collected data on DNA fingerprinted mosquito blood meals to compare how well these two distributions describe the data. Then, we conduct OpenMalaria simulations comparing how assumptions of the distribution of EIR heterogeneity affect relationships between different malaria outcomes.

## 2 Methods

### 2.1 Data analysis

Guelbeogo et al. [26] used DNA fingerprinting of human blood meals (matching blood meal DNA to individuals) from wild-caught mosquitoes in Balonghin (health district of Saponé, Burkina Faso) to understand biting heterogeneity between individual human hosts. Here, malaria transmission occurs seasonally between August and December with high prevalence (>80%) during this season [27]. These surveys where performed at the end of the 2013 transmission season (October–December), and at the start (June – July) and peak (September) of the following season in 2014. Each survey indoor mosquito collections were performed in 20–40 households with at least one household member <15 years of age. For each household mosquitoes were collected between 7 and 9 AM by mouth aspiration from walls and ceilings for 5–7 consecutive days within a survey. The head-thoraces of bloodfed mosquitoes were used to identify their species and infection status by PCR.

Here, we use the data from sporozoite positive mosquitoes caught during the study to analyse four datasets; data collected in each of the three surveys and a combined dataset. We only use data from households included in all three surveys, giving a total of 81 individuals. For each individual host we have the number of sporozoite positive blood-fed mosquitoes which where captured and DNA fingerprinted to the individual.

For each dataset we fit two hierarchical Bayesian models to the rate of infectious bites per individual (*i*); *λ_i_*. We use these Bayesian hierarchical models to consider the variations in the biting rates between individuals. Both models account for variations in biting rates between individuals, but differ in their distributional assumptions: one assumes the individual biting rates follow a lognormal distribution, whilst the other assumes they follow a gamma distribution.

For each model, it is assumed the total number of blood meals found to match each individual follows *Poisson*(*λ_i_*) distribution.

Bayesian inference was performed in Stan [28] in Rstudio [29]. Weakly informed priors were used (Table 1). For each model parameterised, we run four Markov chains with 10,000 iterations, removing the first 5000 for burn-in. The convergence of chains was checked using the diagnostics available within Stan, including the R-statistic and effective sample size.

**Table 1:**
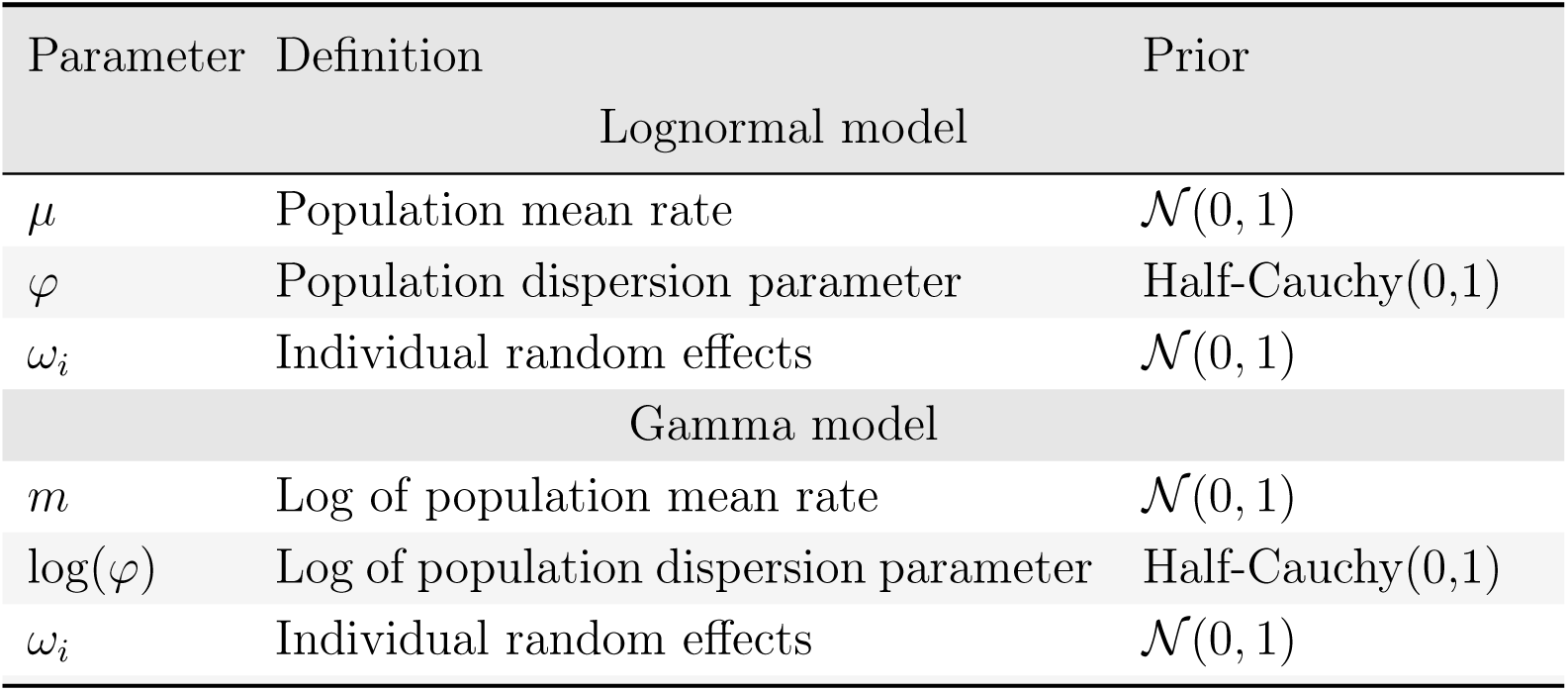
Hyperparamters of the lognormal and gamma models. *i* denotes the individual for parameters that vary between individual hosts.

| Parameter | Definition | Prior |
| --- | --- | --- |
| Lognormal model |  |  |
| $\mu$ | Population mean rate | $\mathcal{N}(0, 1)$ |
| $\varphi$ | Population dispersion parameter | Half-Cauchy(0,1) |
| $\omega_i$ | Individual random effects | $\mathcal{N}(0, 1)$ |
| Gamma model |  |  |
| $m$ | Log of population mean rate | $\mathcal{N}(0, 1)$ |
| $\log(\varphi)$ | Log of population dispersion parameter | Half-Cauchy(0,1) |
| $\omega_i$ | Individual random effects | $\mathcal{N}(0, 1)$ |

To compare models, we estimate the marginal log-likelihoods for each model for each dataset, using the *bridgesampling* package [30], averaging over 10 repetitions of the bridge sampling procedure to obtain an empirical estimate of the estimation uncertainty. We calculate posterior model probabilities (PMPs) from the estimated marginal log-likelihoods to quantify the relative support for each model, where the PMP represents the probability of each model being the true data-generating model, assuming equal prior model probabilities.

#### Lognormal model

For the lognormal model the bite rates (*λ_i_* ∈ *λ*) are considered to follow a lognormal distribution. We consider *ω* which describes how log(*λ*) varies between individuals, with each individual’s log(*λ_i_*) deviating from the mean *m* according to the scale of the standard deviation (*σ*). We denote the individuals biting rates with a subscript *i*, for *i* = 1, 2, . . . , *N* elements, therefore the individuals rates are given as

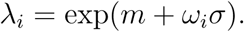

The expected value of the distribution, i.e. the mean of the rates, is therefore

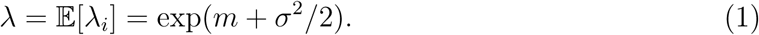

The variance of the rates is

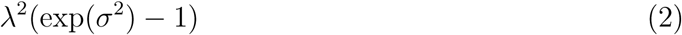

and consequently the standard deviation is

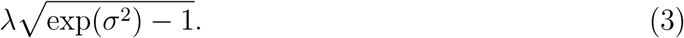

The CV, which provides a standardised measure of dispersion, is given by

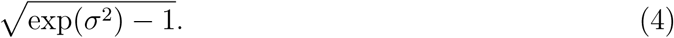

#### Gamma model

For the gamma model, we model the variation in individual biting rates using a Bayesian hierarchical approach with a gamma distribution. The distribution is parameterised by the population mean (*µ*) and dispersion parameter (*φ*). The individual rates (*λ_i_*) are derived from a transformation of standard normal random effects (*ω_i_*) to approximate the gamma distribution:

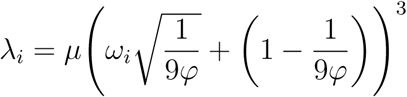

This formulation uses the Wilson-Hilferty transformation to convert standard normal random effects into gamma-distributed values. The *ω_i_* terms are individual random effects that follow a standard normal distribution, and the transformation ensures that the resulting distribution of rates approximates a gamma distribution with mean *µ* and variance *µ*^2^*/φ*.

This formulation has several desirable properties: (i) the population mean is preserved as *µ*, (ii) the variation scales with *µ* (larger means have larger absolute variation) and (iii) the CV is controlled by *φ*, ensuring that as *φ* increases, the variation between individuals decreases, matching the gamma distribution’s inherent variance structure.

This parameterisation can also be expressed in terms of the traditional gamma shape (*α*) and rate (*β*) parameters, where

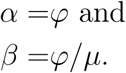

### 2.2 OpenMalaria simulations

OpenMalaria represents a stochastic, individual-based framework for analysing malaria transmission dynamics and evaluating intervention strategies and their public health impact [31–33]. The modelling platform has been applied to forecast the effectiveness of diverse control strategies, including vector-control and pharmaceutical interventions. Within human hosts, OpenMalaria simulates both asexual and sexual parasite stages, immunity acquisition through infection and intervention exposure, drug pharmacokinetics and pharmacodynamics, as well as antimalarial resistance. The model considers clinical disease episodes, both severe and uncomplicated, with individual treatment pathways dictated by a customisable health system architecture. Within OpenMalaria, mosquito dynamics encompass the complete life-cycle and behavioural patterns of malaria-transmitting Anopheles species, incorporating key parameters describing host-seeking behaviour, vector survival and responsiveness to insecticidal interventions.

Transmission intensity in OpenMalaria simulations is parameterised through the EIR, the expected number of infectious bites an adult receives annually. In the absence of heterogeneity and interventions, all adults receive the same expected EIR. Introducing heterogeneity creates variation in individual exposure levels, effectively dividing the population into subgroups across a continuous scale of EIRs. Importantly, despite individual variation, the population-level EIR remains equal to the input parameter specified to OpenMalaria. We use the term ‘mean EIR’ to emphasise this distinction between individual-level and population-level measurements.

To introduce heterogeneity, OpenMalaria assigns each individual an exposure factor sampled from a gamma distribution with mean 1 and a coefficient of variation (CV) which controls the level of heterogeneity. A CV of 0 corresponds to homogeneous transmission. The factor is drawn at model initialisation and birth for each individual. At each time step, the rate which individuals receive bites, *α_h,i_*(*t*), is multiplied by this factor. *α_h,i_*(*t*) itself depends on the age of individuals and their received interventions, such as bed nets, as well as on various characteristics of the mosquito population which are not covered in detail here. The resulting product serves as the rate parameter for a Poisson distribution from which the number of infectious bites the individual receives is randomly sampled each time step. Due to the gamma distribution of the heterogeneity factor, the number of bites for a given age and intervention exposure follow a negative negative binomial distribution.

When the mean EIR is large, with a high level of heterogeneity, the gamma distribution can lead to some extreme cases of exposure for some individuals. For example, at a CV of 2.5, the 99.99% percentile of the heterogeneity factor is approximately be 25. For a mean EIR of 30, this corresponds to at least 710 infectious bites per year for 1 in 10,000 individuals. In order to avoid more extreme values, we truncate the heterogeneity factor at a value of 25 for all simulations. This can result in a change in the mean EIR for CV values larger than 2, with a reduction to 97.5% for a CV of 2.5 and 85% for a CV of 3.5 (Figure S1). Conversely, high levels of heterogeneity will also result in very low heterogeneity factors for some individuals, effectively excluding them from malaria transmission entirely. To assess the proportion of the population at risk, we track the age of each individual’s first malaria infection. If individuals are not infected before the age of 60, we consider them to be excluded from malaria transmission.

We consider the EIR-prevalence and EIR-incidence relationships alongside age-prevalence and age-incidence curves to assess the impact of heterogeneity on these fundamental epidemiological relationships at equilibrium. To ensure equilibrium conditions, each OpenMalaria simulation includes a 100-year burn-in period, allowing the system to reach a stable state where population immunity fully reflects long-term exposure patterns. This burn-in guarantees that all reported results and figures represent true steady-state values, eliminating any transient dynamics that might otherwise confound the analysis. We explore transmission intensities ranging from mean EIRs of 1 to 300, combined with heterogeneity levels parameterised through CVs from 0 to 2.5. For simplicity, we assume no seasonality in malaria transmission. The age distribution of the human population is taken from the parameterisation used to fit the base parameters of OpenMalaria [34] and effective coverage for uncomplicated malaria cases is 37% [35]. A detection threshold of 40 parasites/µL of blood is used for prevalence calculations. The mosquito species is *An. funestus* with a blood index of 0.98.

In a second analysis, we explore how introducing heterogeneity can improve the fit of OpenMalaria to age-incidence and age-prevalence curves derived from field data [36]. The considered curves are used for fitting OpenMalaria parameters [37], but a good fit cannot be achieved for some of the locations under the assumption of homogenous biting exposure. For these locations, there was likely a high degree of heterogeneity, but the extent of heterogeneity is unknown and a method for introducing heterogeneity into OpenMalaria has not been previously established, which is why heterogeneity has not been included in the fitting process. The parameterisation of mosquito bionomics, population structure, interventions, seasonality and many other factors for the scenarios are adapted to each site of data collection [37]. We run all the scenarios used in the fitting process of these curves including those where we do not necessarily suspect hetereogeneity to be present. We vary the CV parameter to determine the level of heterogeneity that minimises the sum of squared distances from model output to the data. The optimal CV value is determined independently for the age-prevalence and age-incidence curves for each scenario. This allows us to assess the agreement of the estimated heterogeneity between the two curves for a given location. To quantify the magnitude of change introduced by heterogeneity, we calculate the sum of squared differences between model outputs under homogeneous transmission and those with the best-fit CV values.

## 3 Results

### 3.1 Data analysis

The marginal log-likelihood estimates consistently favoured the gamma model across all temporal subsets of the data, as well as the combined dataset (Table 2). The estimated values of the marginal log-likelihoods varied substantially across the temporal subsets, with the peak period showing markedly higher values (76.679 and 83.766 for lognormal and gamma models, respectively) compared to the start and end periods. This pattern suggests better model fit during the peak period.

**Table 2:** The median and interquartile range width of the marginal log-likelihood estimates for the lognormal and gamma models, as well as the mean posterior model probabilities (PMPs). Results are given to 3 decimal places.

| Dataset | Model | Median | Interquartile range width | PMPs |
| --- | --- | --- | --- | --- |
| Start | Lognormal | 40.973 | 0.011 | 0.000 |
|  | Gamma | 49.917 | 0.018 | 1.000 |
| Peak | Lognormal | 76.679 | 0.014 | 0.001 |
|  | Gamma | 83.766 | 0.026 | 0.999 |
| End | Lognormal | 42.732 | 0.017 | 0.003 |
|  | Gamma | 48.512 | 0.015 | 0.997 |
| Combined | Lognormal | 113.501 | 0.022 | 0.006 |
|  | Gamma | 118.594 | 0.017 | 0.994 |

The precision of the maginal log-likelihood estimates, as measured by the interquartile range, ranged from 0.011 to 0.026. There was no consistent pattern in which the model showed a lower estimation uncertainty.

The PMPs provide a direct measure of the relative support for each model in the data. The PMPs showed a consistent pattern, with the gamma model achieving probabilities between 0.994 and 1 in all datasets, indicating stronger support for this model specification.

Figure 1 shows that the fitted gamma distribution describes the data better than the log-normal. The composite estimates for the mean and CV of the distributions are much more uncertain for the lognormal distribution (Figure S2). The posterior median for CV ranged from 5.44–12.18 and 1.72–2.14 under the lognormal and gamma distribution assumptions, respectively. For the gamma distribution the CV is lower for the start and end of the seasons, compared to the peak.

**Figure 1:**
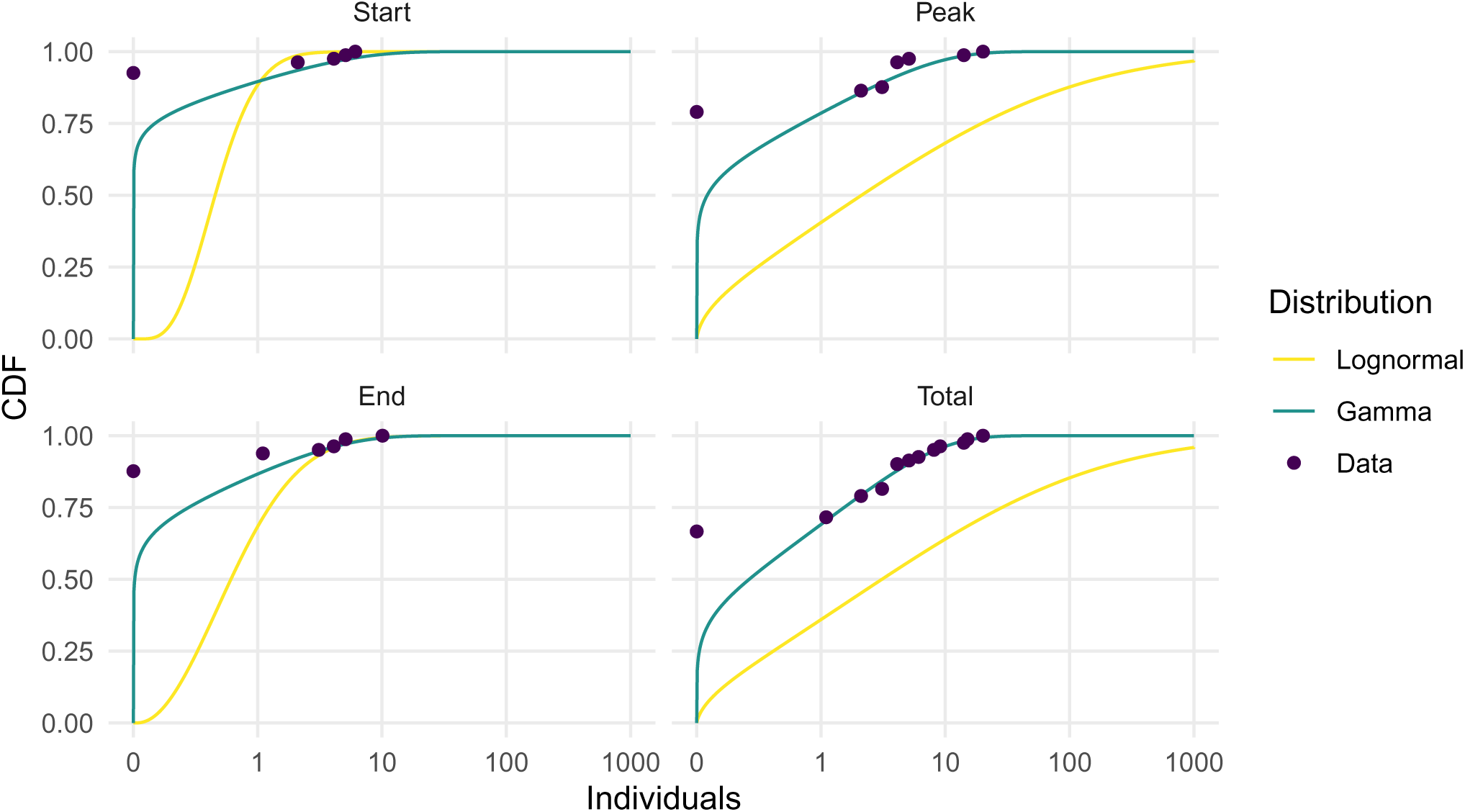
The cumulative density function of mosquito exposure over the human individuals of the fitted lognormal and gamma models and the empirical data.

The lognormal distribution notably underestimates the proportion of individuals with low exposure values, particularly during peak transmission and when considering the combined dataset. This is because the lognormal distribution’s heavier tail forces a slower rise in the CDF to accommodate even a few individuals with higher exposure, sacrificing fit quality for the majority of the population. This sensitivity to outliers is clearly demonstrated when comparing the ’Start’ and ’End’ periods: despite similar exposure patterns for most individuals (fewer than 5 bites), the presence of a single individual receiving 10 bites during the ’End’ period causes a noticeable downward shift in the lognormal CDF. In contrast, the gamma distribution maintains a better balanced fit across the entire range of the data even when accommodating occasional outliers. This robustness to extreme values whilst preserving accuracy for the majority of observations confirms our model selection results that consistently favoured the gamma distribution across all temporal periods.

### 3.2 Effects of heterogeneity at population level

In the absence of heterogeneity, OpenMalaria estimates for malaria prevalence and incidence at the population level reach saturation at relatively low transmission intensities (Figure 2A and 3A). Parasite prevalence peaks at 59% with a mean EIR of 50, with the 50% threshold crossed at a substantially smaller mean EIR of 10. Clinical incidence follows a different pattern, reaching its maximum of 1.25 cases per person per year at an EIR of 9, before declining as the mean EIR increases further. This post-peak reduction occurs because increased exposure strengthens population immunity, thereby reducing clinical episodes. Importantly, this declining pattern is not observed for prevalence, as immunity, whilst reducing parasitaemia, does not eliminate it completely.

**Figure 2:**
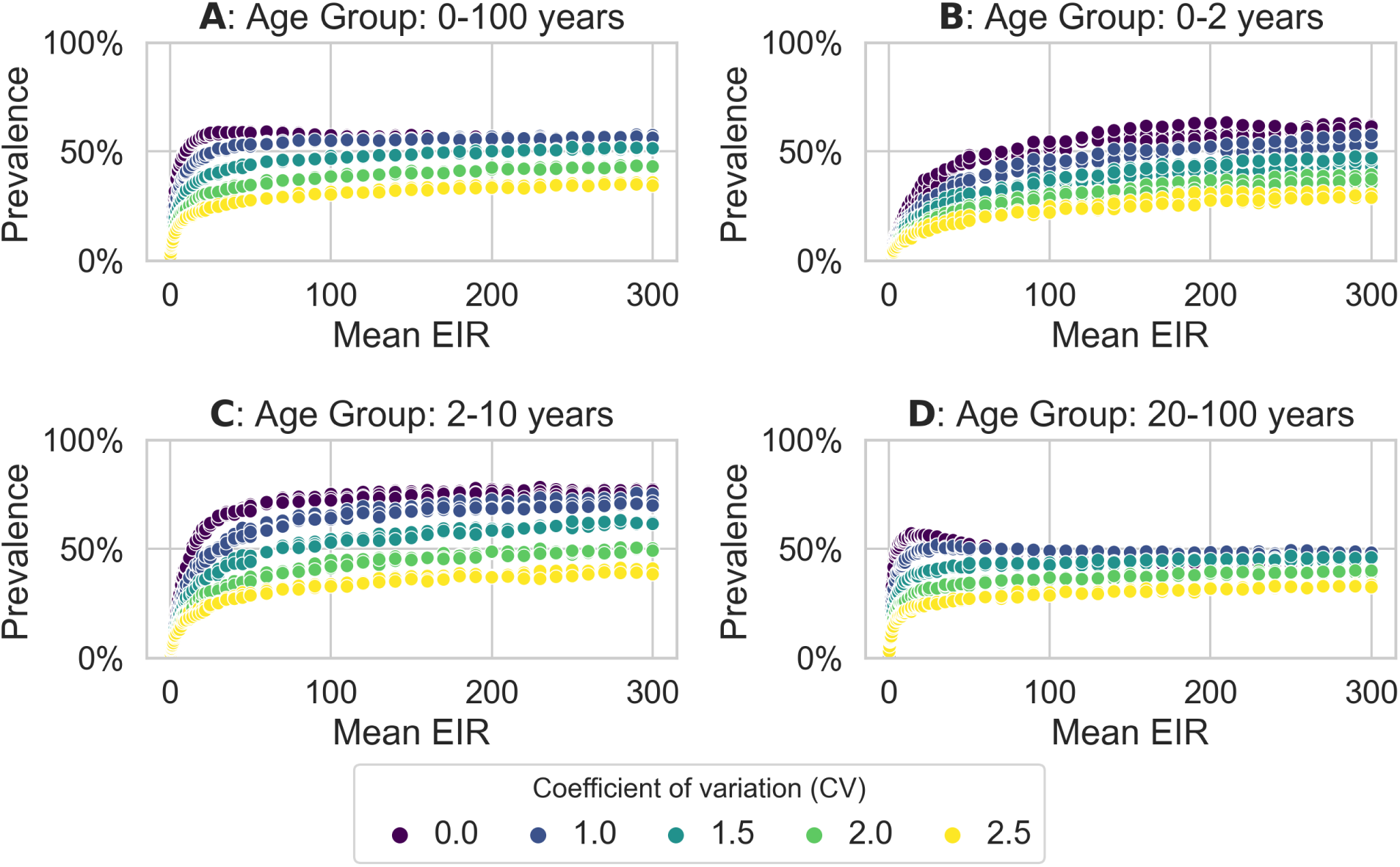
Parasite prevalence across the entire population (A) and specific age groups (B-D) at varying transmission intensities (mean EIRs). Each point represents a simulation. The CV=0 curves can be interpreted as approximations of the EIR-prevalence relationships at the individual level.

**Figure 3:**
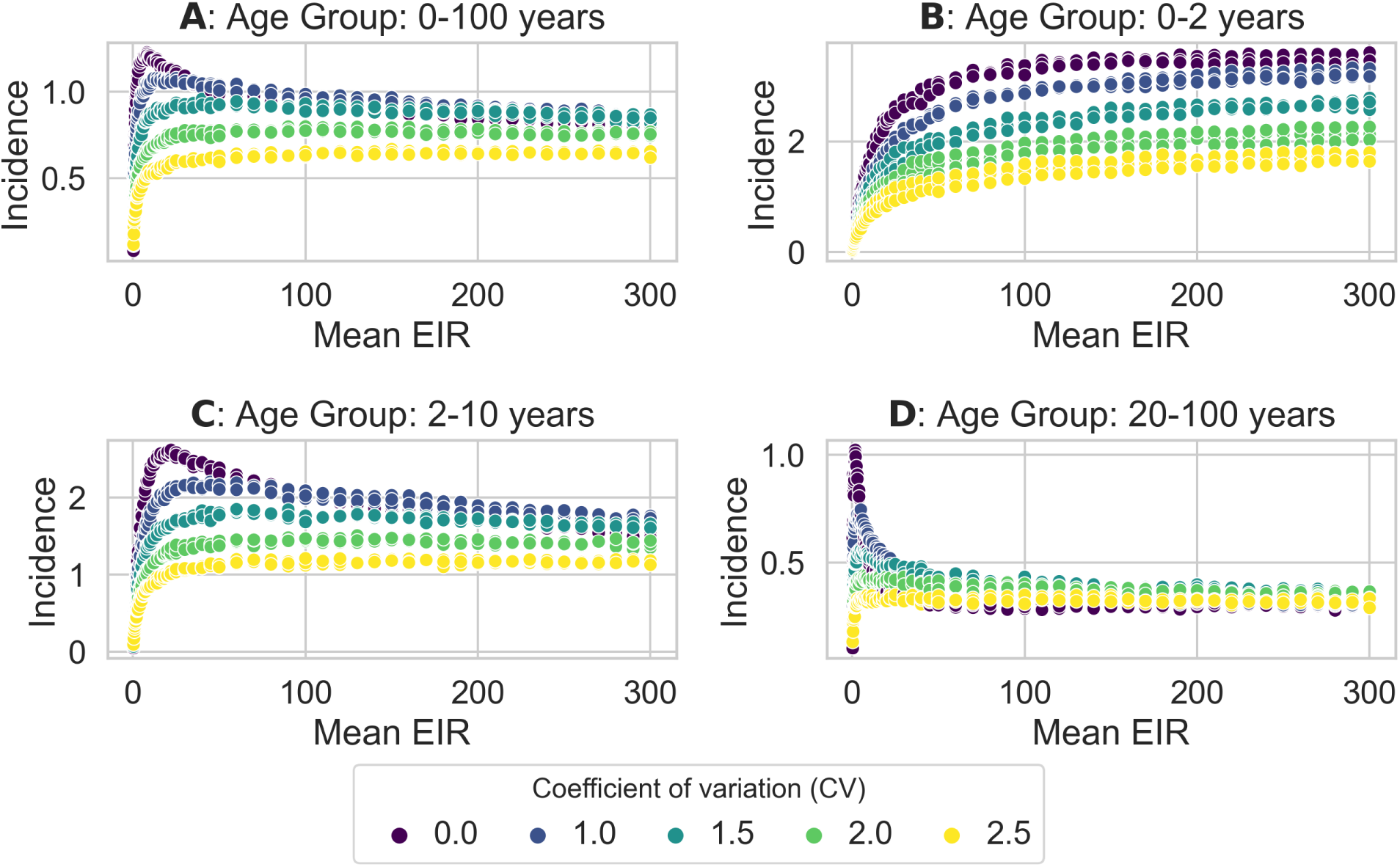
Clinical incidence across the entire population (A) and specific age groups (B-D) at varying transmission intensities (mean EIRs). Each point represents a simulation. The CV=0 curves can be interpreted as approximations of the EIR-prevalence relationships at the individual level.

When introducing heterogeneity with the gamma distribution, more individuals experience an EIR below the mean than above it due to the distribution’s right-skewed nature. As an example, we compare prevalence across all ages between a homogeneous scenario (CV=0) and a heterogeneous scenario (CV=1) at a mean EIR of 100 (Figure S3). As the majority of individuals are shifted below the mean EIR of 100, they experience individual EIRs that result in similar or lower prevalence levels. Whereas, individuals shifted above the mean EIR of 100 experience only marginally higher prevalence. Overall, since there are more individuals with an EIR lower than the mean, this yields lower overall prevalence compared to homogeneous exposure. This effect holds for any mean EIR and increases with heterogeneity, as more individuals experience low EIRs with an increase in CV. This results in prevalence remaining below 40% even at mean EIRs as high as 300 under highest studied heterogeneity conditions (Figure 2).

At the population level, more heterogeneity also results in lower clinical incidence, with a notable exception at low levels of heterogeneity. As an example, considering a mean EIR of 100 and CV = 1 (Figure S4), the redistribution of individuals below the mean EIR drives them towards higher incidence levels due to reduced immunity development. Through this mechanism, low heterogeneity produces slightly higher overall incidence values in high transmission settings (Figure 3A). However, with higher heterogeneity levels, enough individuals are driven towards sufficiently low EIRs that the effect of reduced exposure outweighs the impact of reduced immunity.

### 3.3 Effects of heterogeneity across age groups

The EIR-prevalence and EIR-incidence curves show very different relationships across age groups, both in homogeneous and heterogeneous transmission scenarios. In the absence of heterogeneity, adults exhibit the most distinctive pattern (Figure 3D), with clinical incidence sharply peaking at a low mean EIR of 1.75, before rapidly declining as the mean EIR increases. This pattern reflects immunity dynamics; higher transmission intensity leads to greater accumulated immunity in adults, subsequently reducing clinical episodes. In contrast, younger age groups display increasingly monotonic relationships (Figure 3B-C), where higher mean EIRs consistently correspond to increased clinical incidence. Prevalence exhibits similar age-dependent patterns, with younger groups showing trends toward monotonicity (Figure 2B-D). As the level of heterogeneity is increased, both prevalence and incidence relationships increasingly shift toward monotonicity regardless of age.

As demonstrated in Section 3.2, both overall incidence and prevalence generally decrease with increasing heterogeneity across all mean EIR values. Figure 4A shows that the higher overall incidence observed in settings with lower heterogeneity accumulates primarily in younger age groups, while incidence rates among older individuals remain similar regardless of heterogeneity level. Under homogeneous transmission, most individuals experience their first infections at a young age, acquiring immunity accordingly. As mean EIR increases, these infections occur progressively earlier in life, causing age-incidence curves to decline more steeply with age. For prevalence, lower heterogeneity consistently produces higher estimates across most age groups, with an exception occurring in older individuals exposed to high transmission intensity under low heterogeneity conditions (Figure 4B). This contrasting pattern between prevalence and incidence can be attributed to acquired immunity, which prevents clinical episodes but not infections.

**Figure 4:**
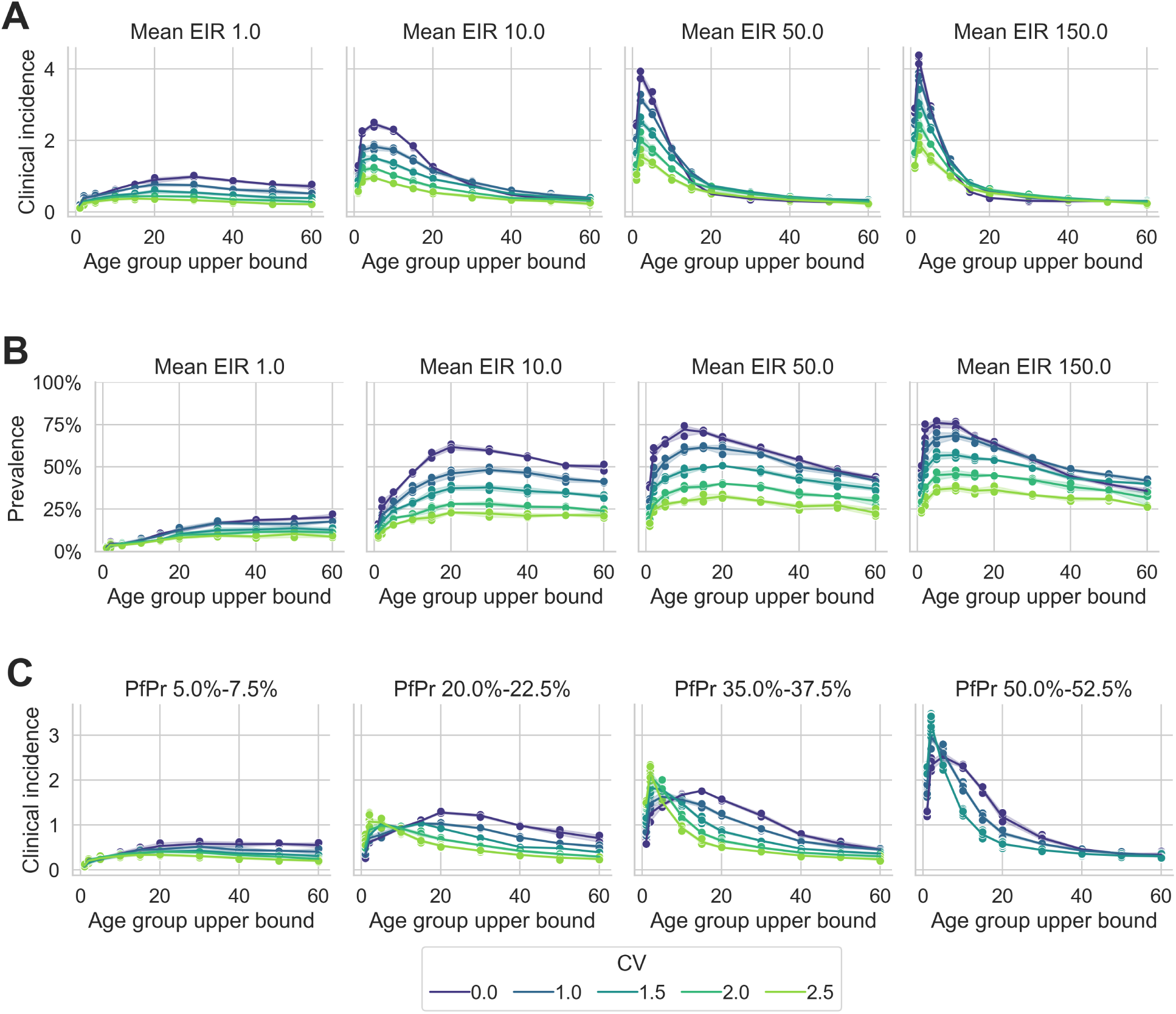
Age-incidence (A) and age-prevalence curves (B) at varying transmission intensities (mean EIRs), and age-incidence curves for varying levels of prevalence (C).

When comparing the age-incidence curves at a given prevalence instead of a given mean EIR, disease burden shifts towards younger age groups as heterogeneity increases (Figure 4C). This pattern emerges because achieving equivalent population-level prevalence requires significantly higher mean EIRs under greater heterogeneity. For example, an overall prevalence of 35-37.5% requires a mean EIR of 4 at CV=0, increasing to a mean EIR of 7 at CV=1, and rising substantially to mean EIRs of 69 and 275 at CV=2 and CV=2.5, respectively. Under homogeneous transmission (CV=0), a relatively low mean EIR results in infrequent infections across the entire population, producing gradual immunity development and allowing infections to occur across a broader age range. In contrast, at high heterogeneity (CV=2.5), the high mean EIR concentrates exposure in a subpopulation that rapidly accumulates infections at an early age, quickly developing immunity and experiencing fewer infections as they age. This pattern — shifting incidence towards younger ages with increasing heterogeneity — persists even when accounting for the effective exclusion of part of the population from infection risk under high heterogeneity conditions (Figures S5 to S7).

For high levels of heterogeneity (CV≥ 2), the population at risk is below 50% for low mean EIRs, gradually increasing to approximately 80% at higher mean EIRs, reaching saturation. In contrast, for lower levels of heterogeneity, the entire population is at risk of infection. This leads to an increase in incidence and prevalence for any given mean EIR when adjusting for the proportion of the population at risk. The age-incidence curves corrected for this effect exhibit the same pattern observed in uncorrected analyses (Figures S5 to S7): higher heterogeneity consistently shifts disease burden towards younger age groups. This demonstrates that the observed pattern is not just a consequence of restricting transmission to a subpopulation, but represents an effect of heterogeneity *within* the malaria-affected subpopulation. In contrast to the uncorrected version, prevalence levels of 65% and higher can be achieved when considering only the population at risk.

### 3.4 Parametrising heterogeneity from age-prevalence and age-incidence curves

For several tested scenarios, incorporating heterogeneity substantially improves the alignment between simulated age-incidence curves and field observations. Figure 5 shows the three scenarios where including heterogeneity the yielded the most improvements. For these scenarios, the sum of squared distances — measuring deviation between observed data and model predictions — decreased by approximately 95%. The optimal heterogeneity levels varied considerably: the scenarios from Manhica and Matola (both Mozambique) required very high heterogeneity, while the scenario from Koundou, Cameroon showed best results with more moderate heterogeneity. For prevalence, the best-fit CV values was substantially smaller with a much more moderate improvement in the fit, with a CV of 0 producing the best fit for scenario Manhica. For all scenarios tested, Figures S8 and S9 show model outputs when the model is parameterised using the CV value which produced the best fit and Table S1 details the improvement by introducing heterogeneity.

**Figure 5:**
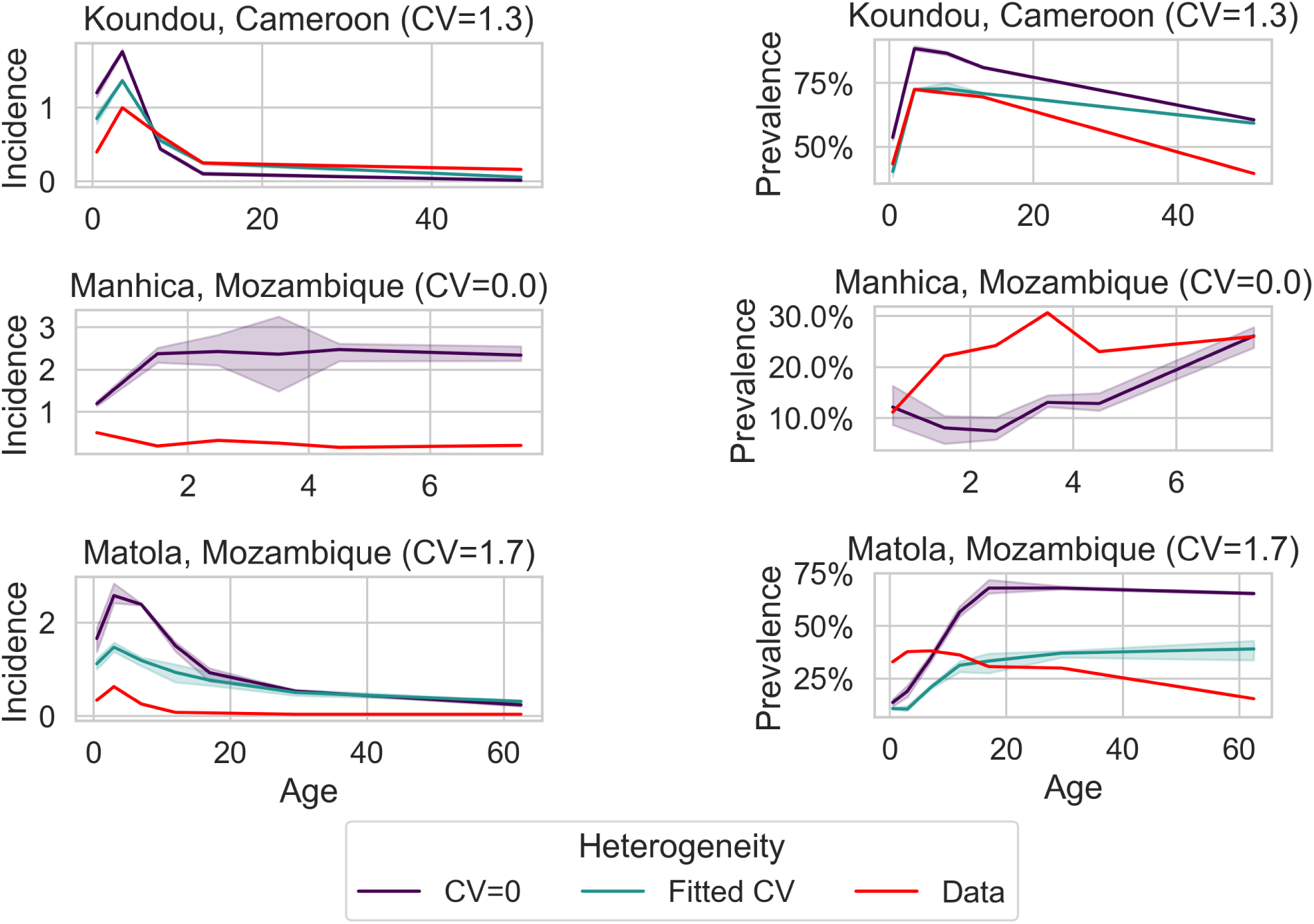
Age-incidence (left column) and age-prevalence curve (right column) of three scenarios that were most improved by added heterogeneity. Fitted CV curves show predictions with the model parameterised using the best-fit CV value, which is also given in the brackets.

## 4 Discussion

This study advances our understanding of heterogeneity in malaria transmission beyond the foundational theoretical work of Dye and Hasibeder [23]. Whilst they established that heterogeneous biting increases the basic reproductive rate and demonstrated this effect using field data, our work provides a rigorous statistical framework for determining which probability distributions best characterize this heterogeneity. Integrating heterogenous biting exposure into OpenMalaria leads to substantial changes in basic equilibrium epidemiological relationships.

Within OpenMalaria, the effects of heterogeneity are driven by two interrelated mechanisms: saturation effects and immunity development. Saturation effects arise when individuals receive multiple infectious bites. Beyond a certain threshold, additional bites contribute little to further disease incidence. If an individual is already experiencing a clinical malaria episode, subsequent infections progressing to the blood stage are not counted as separate episodes. Thus, whilst the risk of developing an infection increases with the number of infectious bites, each additional bite has progressively less impact on disease risk.

Immunity effects add further complexity to transmission dynamics. At equilibrium, where individuals receive consistent average exposure over extended periods, those exposed to higher bite rates develop stronger immunity. Consequently, increasing bite exposure for an individual typically results in fewer clinical episodes over time, particularly in adults where immunity has accumulated over years. This creates an inverse relationship between accumulated exposure and infection risk.

The interaction between saturation and immunity explains the observed effects of heterogeneity in OpenMalaria simulations. Generally, higher heterogeneity reduces population-level incidence due to saturation effects amongst heavily exposed individuals. However, immunity can reverse this effect in adults for certain levels of heterogeneity. Under homogeneous exposure, most adults develop sufficient immunity to substantially reduce disease incidence. With heterogeneity, whilst individuals with high exposure also develop strong immunity, some adults remain relatively unexposed and fail to develop adequate immunity for disease prevention. These individuals experience occasional infections throughout adult life, thereby increasing overall adult incidence despite the general trend towards lower population-level transmission.

We explored the effect of heterogeneity over a wide range of transmission settings, including those with very high EIR. Previous research has shown greater heterogeneity in lower EIR settings [16, 38], with pockets of infections. Whilst including the combination of high transmission activity and high heterogeneity in the analysis provides a comprehensive assessment of the effects of heterogeneity in OpenMalaria, such settings are less likely to occur in natural contexts. We can implement decreasing heterogeneity with increasing EIR in Openmalaria, but currently have insufficient data to parameterise this relationship appropriately.

The analysis of the heterogeneity in human blood meal data across the three survey periods showed that this inverse correlation between EIR and heterogeneity may not only be applicable regionally, but also through seasonal variations in transmission. A larger CV was observed during the start and end of the season, when transmission levels were lower. This temporal variation in heterogeneity, with larger CVs during periods of lower transmission, adds an important temporal dimension to our understanding of exposure patterns that was not captured in earlier theoretical frameworks. We, however, did not include seasonality in the simulations to avoid additional interactions and isolate the effects of heterogeneity in exposure between individuals. However, seasonality is found in most malaria transmission settings and future work could examine how these temporal patterns in the degree of individual-level heterogeneity affect transmission dynamics.

By comparing gamma and lognormal distributions using Bayesian hierarchical models and modern model selection techniques, we show that the gamma distribution better captures the observed patterns of exposure heterogeneity. Whilst the lognormal distribution has theoretical appeal for modelling extreme values through its heavy tail, it systematically overestimates the proportion of individuals receiving very high exposure whilst underestimating those with low exposure for our datasets. Although the gamma distribution lacks the heavy tail of the lognormal, it provides a more accurate representation of the actual distribution of biting exposure in the population.

The absence of heavy tails in the gamma distribution has important consequences for modelling saturation and immunity effects. Without the extreme outliers characteristic of heavy-tailed distributions, fewer individuals reach the saturation threshold where additional bites yield diminishing returns on disease incidence. Simultaneously, the gamma distribution’s more moderate extreme values mean that highly exposed individuals accumulate immunity more gradually, reducing the high contrast between high and low exposure groups. This modulation of both saturation and immunity effects likely produces more realistic transmission dynamics compared to the polarised populations generated by heavy-tailed distributions. Howewer, even after right-truncation, the gamma distribution can result in some extreme differences in exposure between individuals under high heterogeneity. Possible approaches to this problem are additional left-truncation setting a minimal exposure or alternative distributions that lead to less extreme values, for example a triangular distribution or a non-parametric leading to a 80/20 distribution of biting exposure.

Many studies that estimate EIRs report averages and confidence intervals over the duration of the study. More detailed data for an individual level would help further our understanding of these distributions. To understand heterogeneity at the individual level, studies that perform human landing catches or DNA fingerprinted of resting, blood-fed mosquitoes would be the most informative. While DNA fingerprinting of mosquito blood meals provides valuable insights into biting heterogeneity, potential sampling biases warrant consideration. Our analytical approach assumes that the absence of matched blood meals for certain individuals genuinely reflects lower biting rates. However, methodological limitations could artificially inflate heterogeneity estimates if mosquitoes that fed on particular individuals are systematically underrepresented in collections. This might occur due to spatial variation in mosquito resting behaviours, household-specific factors affecting mosquito collection efficiency, or stochastic sampling effects in households with few mosquitoes.

In this study we demonstrated that individual-level heterogeneity can be directly parameterised from data. We also showed that heterogeneity can allow the model to fit age-incidence curves from field data that could not be reproduced under homogeneous exposure. Whilst heterogeneity was documented during data collection from studies in Manhica, Mozambique and Dielmo, Senegal (scenarios s30 and s32 respectively in Table S1), this was not the case for all scenarios showing improved fit. Importantly, heterogeneity primarily improves the fit by lowering simulated incidence levels, though such reductions could also arise from factors such as incomplete case reporting. Fitting to age-prevalence curves yielded markedly different results: heterogeneity estimates were substantially lower and improvements were minimal. Additionally, there was no statistically significant correlation between the heterogeneity fitted to age-prevalence and age-incidence curves (Table S1). This could be due to two possible reasons. First, this inconsistency might suggest that heterogeneity is not the primary driver of the observed age-specific patterns in both indicators. Alternatively, the discrepancy could reflect fundamental differences in how these indicators are measured: prevalence data is typically obtained from cross-sectional surveys, whilst incidence relies on active surveillance systems. These measurement approaches are subject to different biases, possibly skewing results in opposing directions.

The higher heterogeneity observed during periods of lower transmission suggests that control interventions may face greater challenges in elimination settings than previously recognised. The incorporation of appropriate heterogeneity distributions in transmission models enhances their appropriateness for modelling transmission as we approach elimination settings. Hasibeder and Dye [39] examined how spatial clustering of vectors and hosts could further amplify transmission beyond heterogeneity. Their mathematical analysis showed that transmission dynamics could be dramatically altered by strong associations between particular groups of mosquitoes and hosts. A route of future studies is the explicit inclusion of spatial heterogeneity in OpenMalaria with distinct mosquito populations.

Not considering heterogeneity in transmission when evaluating control tools can lead to inaccurate estimated impact [3, 11]. If control programmes fail to reach individuals with relatively high exposure compared to the majority of the population, they are likely to be much less effective. Future work should explore methods for identifying and targeting high-exposure individuals in elimination settings. This approach would focus interventions where they will have the greatest effect on reducing transmission, maximising the impact of limited resources.

## Data Availability

No data was produced. Data sources are cited in the manuscript.

## Acknowledgements

Calculations were performed at sciCORE (http://scicore.unibas.ch/) scientific computing center at University of Basel.

## Ethical approval

The authors confirm that the ethical policies of the journal, as noted on the journal’s author guidelines page, have been adhered to. No ethical approval was required.

## Author contributions

**LK:** Conceptualisation, Methodology, Software, Validation, Formal analysis, Investigation, Writing - Original draft, Writing - Review & Editing, Visualisation **AC:** Conceptualisation, Methodology, Software **MAP:** Conceptualisation, Resources, Funding acquisition **NC:** Conceptualisation, Methodology, Resources, Writing - Review & Editing, Supervision, Funding acquisition **ELF:** Conceptualisation, Methodology, Software, Validation, Formal analysis, Investigation, Data curation, Writing - Original draft, Writing - Review & Editing, Visualisation, Project administration, Funding acquisition

