## Supplementary Figure for "Exploring heterogeneity in mosquito exposure and attraction and its implications for malaria transmission"

### Supplementary figures and tables

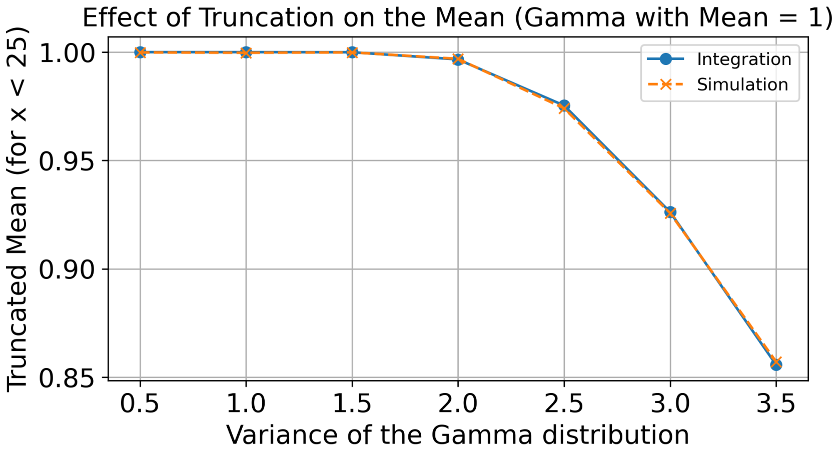


Figure S1: Effect on the mean of truncating a gamma distribution with original mean 1 at a truncation value of 25. Up to a coefficient of variation (CV) of 2, the effect on the mean is limited. At higher CVs, the mean can be substantially altered due to truncation. The values were calculated once using numerical integration of the gamma probability density function and once through random number draws to ensure robustness of the results.

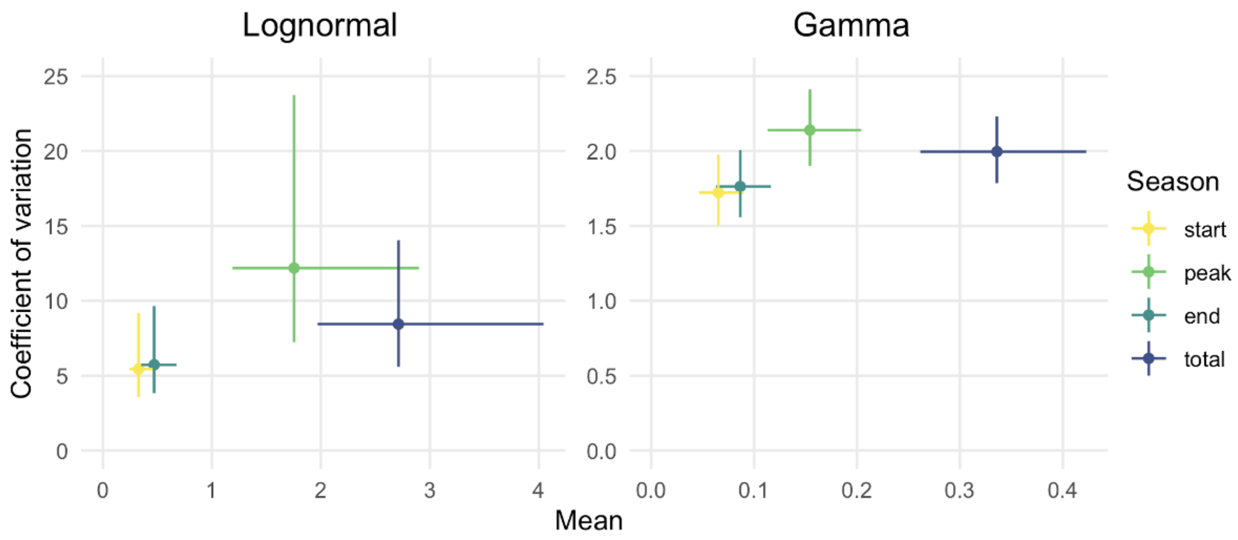


Figure S2: Parameter estimate median (dot) and interquartile range (line) for each model and dataset.

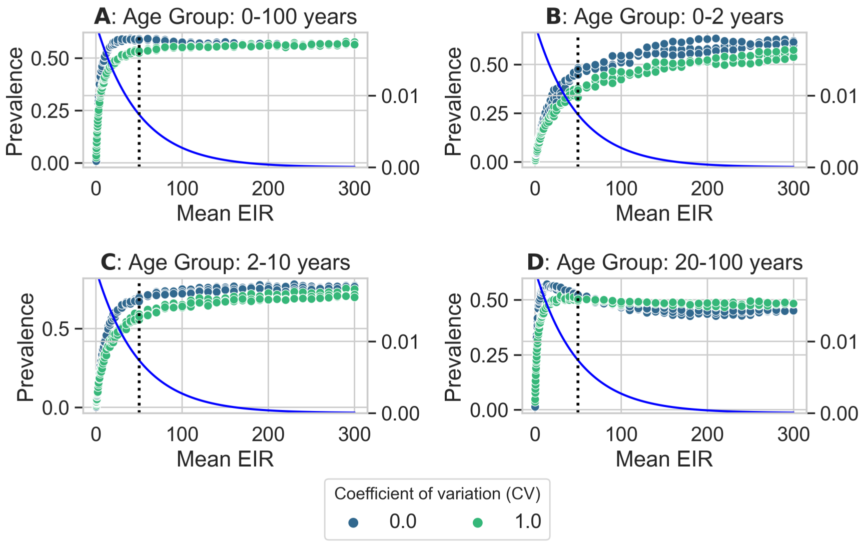


Figure S3: Prevalence of clinical malaria cases across the entire population (A) and specific age groups (B-D) at varying transmission intensities (mean EIRs). Each point represents a simulation. The blue line represents the distribution of EIRs of individuals at a mean EIR of 100 (black dotted line) and a CV of 1.0. The CV=0 curves can be interpreted as approximations of the EIR-prevalence relationships at the individual level.

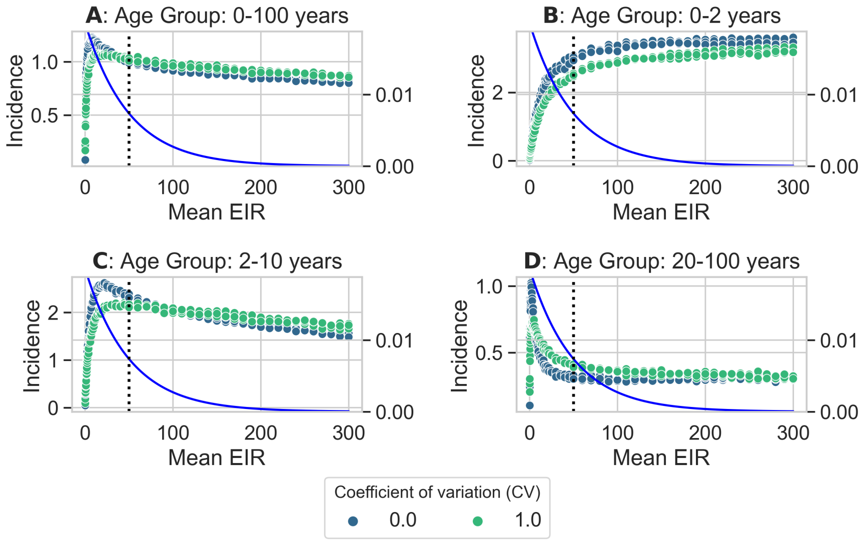


Figure S4: Clinical malaria incidence across the entire population (A) and specific age groups (B-D) at varying transmission intensities (mean EIRs). Each point represents a simulation. The blue line represents the distribution of EIRs of individuals at a mean EIR of 100 (black dotted line) and a CV of 1.0. The CV=0 curves can be interpreted as approximations of the EIR-prevalence relationships at the individual level.

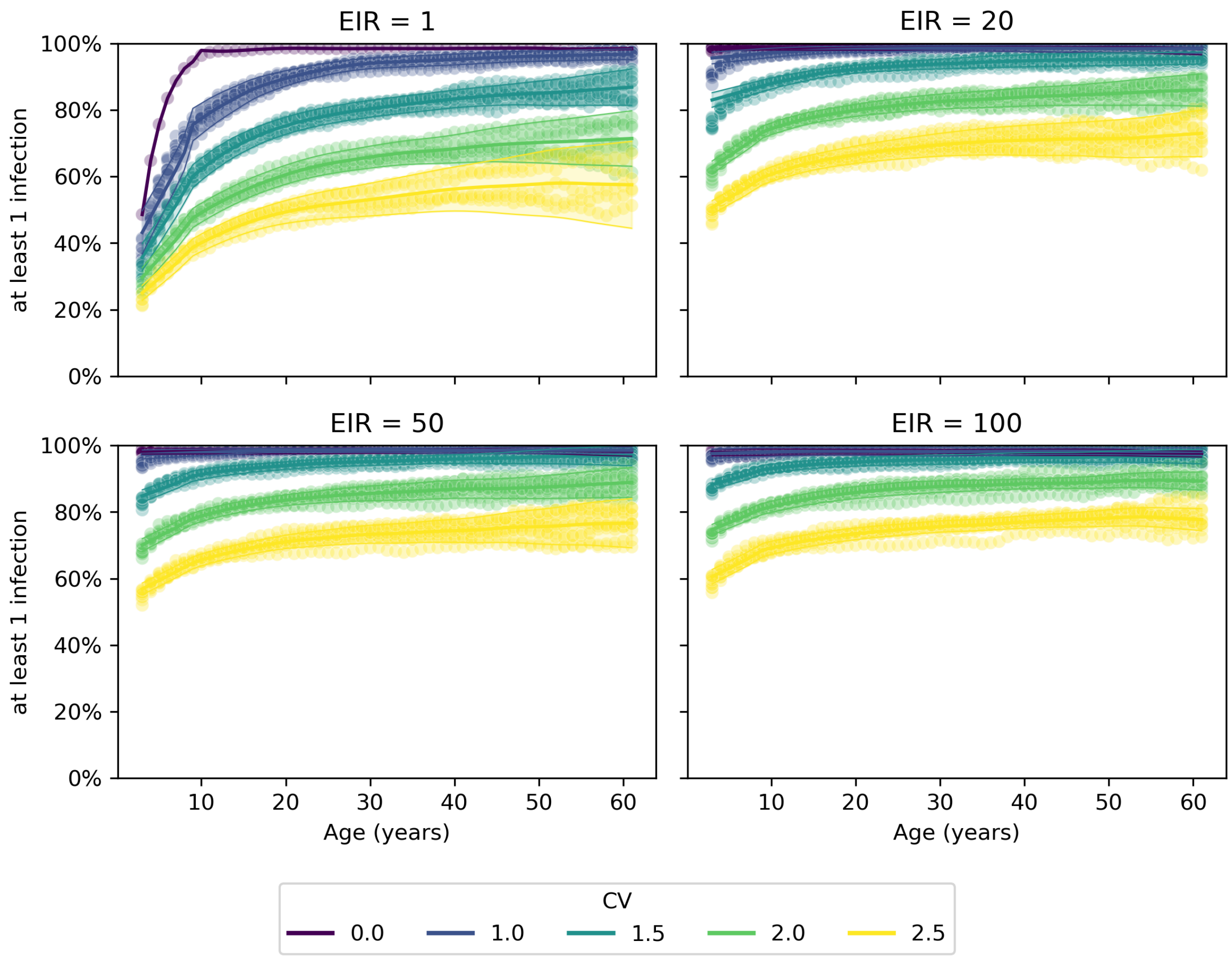


Figure S5: Proportion of individuals which have experienced their first infection at each age. This fraction can be non-monotonous due to stochastic migration effects contained in OpenMalaria to keep the age distribution constant.

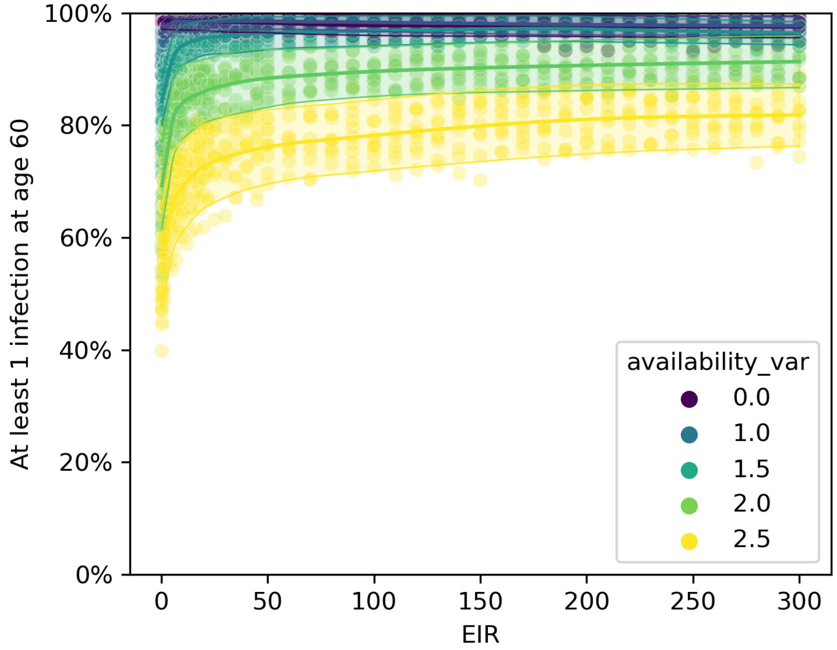


Figure S6: Proportion of individuals that experienced at least one infection at age 60, used as a proxy for the proportion of the population at risk. This corresponds to the last data point for each EIR as plotted in Figure S5.

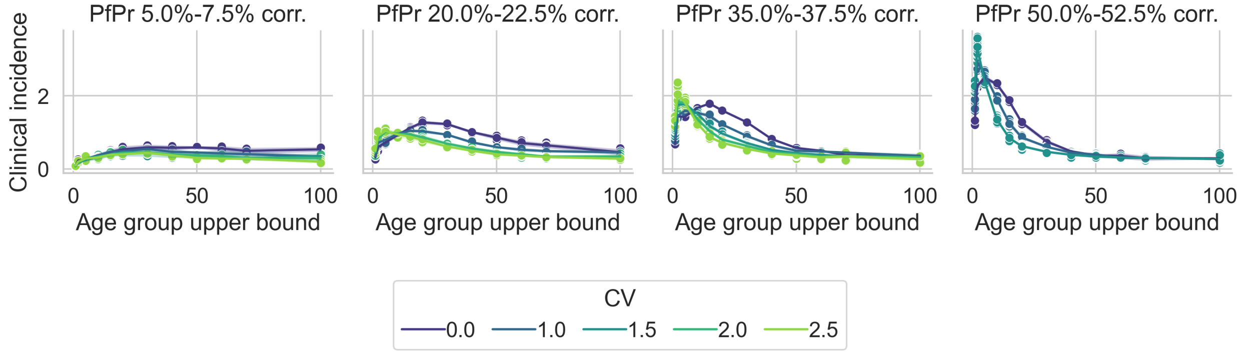


Figure S7: Age-incidence curve over prevalences corrected for the fact that under high levels of heterogeneity, some individuals do not experience infections over their lifetimes. The incidence and the prevalence from the overall population were divided by the proportion of the population at risk.

| | | Incidence | | Prevalence | |
|---|---|---|---|---|---|
| Site | Location | Best-fit CV | Improvement | Best-fit CV | Improvement |
| s01 | Ndiop, Senegal | 0.4 | 32.2% | 1.4 | 16.1% |
| s02 | Asembo Bay, Kenya | 0.1 | 4.7% | 1.0 | 1.6% |
| s03 | Ebolakounou, Cameroon | 0.5 | 6.9% | 1.0 | 3.8% |
| s04 | Koundou, Cameroon | 1.9 | 93.1% | 1.3 | 6.2% |
| s09 | Dakar, Senegal | 1.3 | 10.5% | 0.0 | 0.0% |
| s16 | Katiola, Côte d'Ivoire | 0.0 | 0.0% | 0.0 | 0.0% |
| s17 | Korhogo "R1", Côte d'Ivoire | 2.8 | 69.8% | 0.0 | 0.0% |
| s18 | Korhogo "R2", Côte d'Ivoire | 0.6 | 1.8% | 0.0 | 0.0% |
| s30 | Manhica, Mozambique | 3.3 | 93.6% | 0.0 | 0.0% |
| s32 | Matola, Mozambique | 3.4 | 96.7% | 1.7 | 3.3% |
| s34 | Dielmo, Senegal | 0.2 | 6.0% | 0.0 | 0.0% |

Table S1: Fitted heterogeneity to age-incidence curves (middle section of the table) and age-prevalence curves (right section of the table). The improvement columns show the reduction in the sum squared error between the curves without heterogeneity and the curves with fitted heterogeneity. If the fitted CV equals 0 (i.e. no heterogeneity), the improvement is 0%. While at least some heterogeneity is fitted for most incidence curves, including substantial heterogeneity for scenarios s4, s30, and s32, much smaller values of heterogeneity were fitted for the age-prevalence curves. The fitted CV for incidence and prevalence has a weak, nonsignificant correlation of 0.18. Spearman rank correlation is also nonsigfnificant.

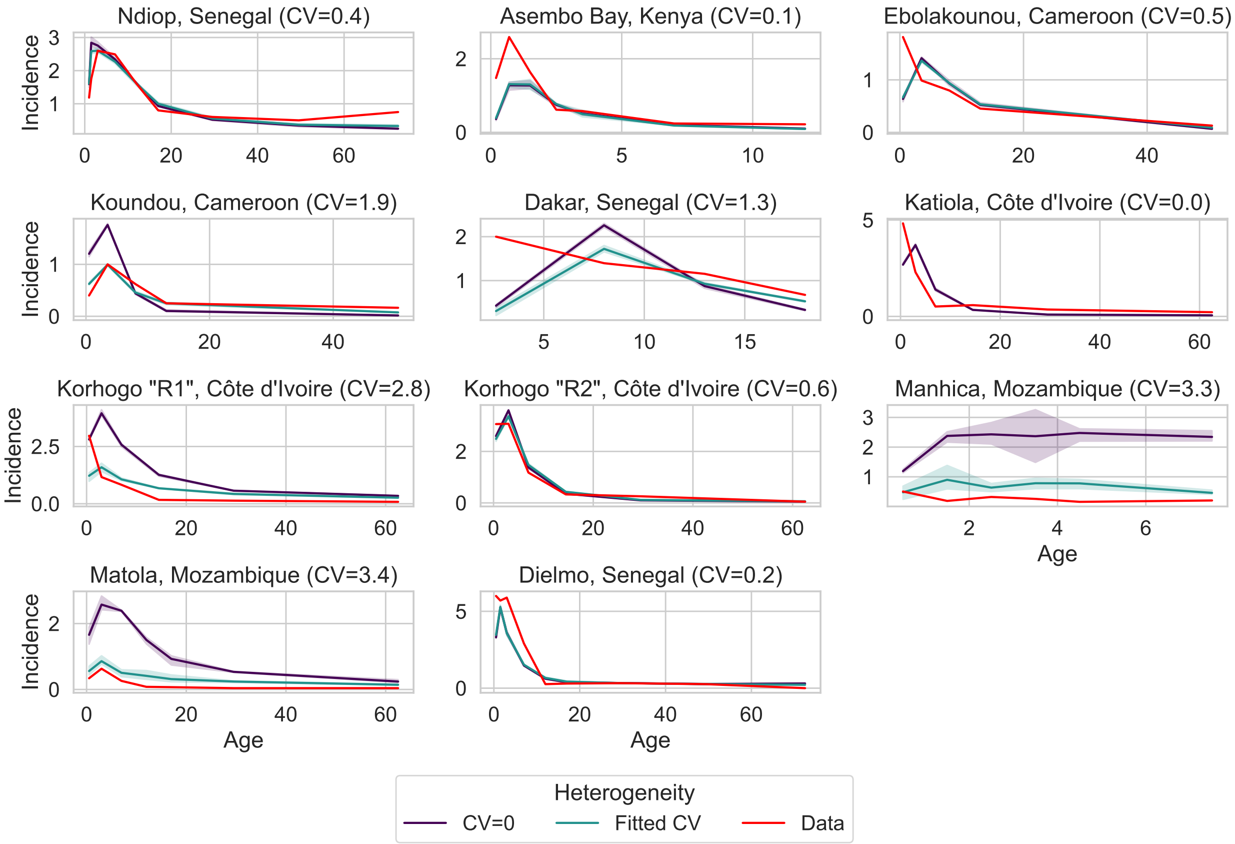


Figure S8: Age-incidence curves collected by Battle et al. [1] used for the fitting of Open-Malaria. Heterogeneity was fitted through the coefficient of variation (CV) such that the distance between data and the curve generated by OpenMalaria was minimized as measured by sum of squares. The fitted CV value is given in brackets behind the scenario names. The improvement of the fit to the data with heterogeneity compared to no heterogeneity is given in Table S1.

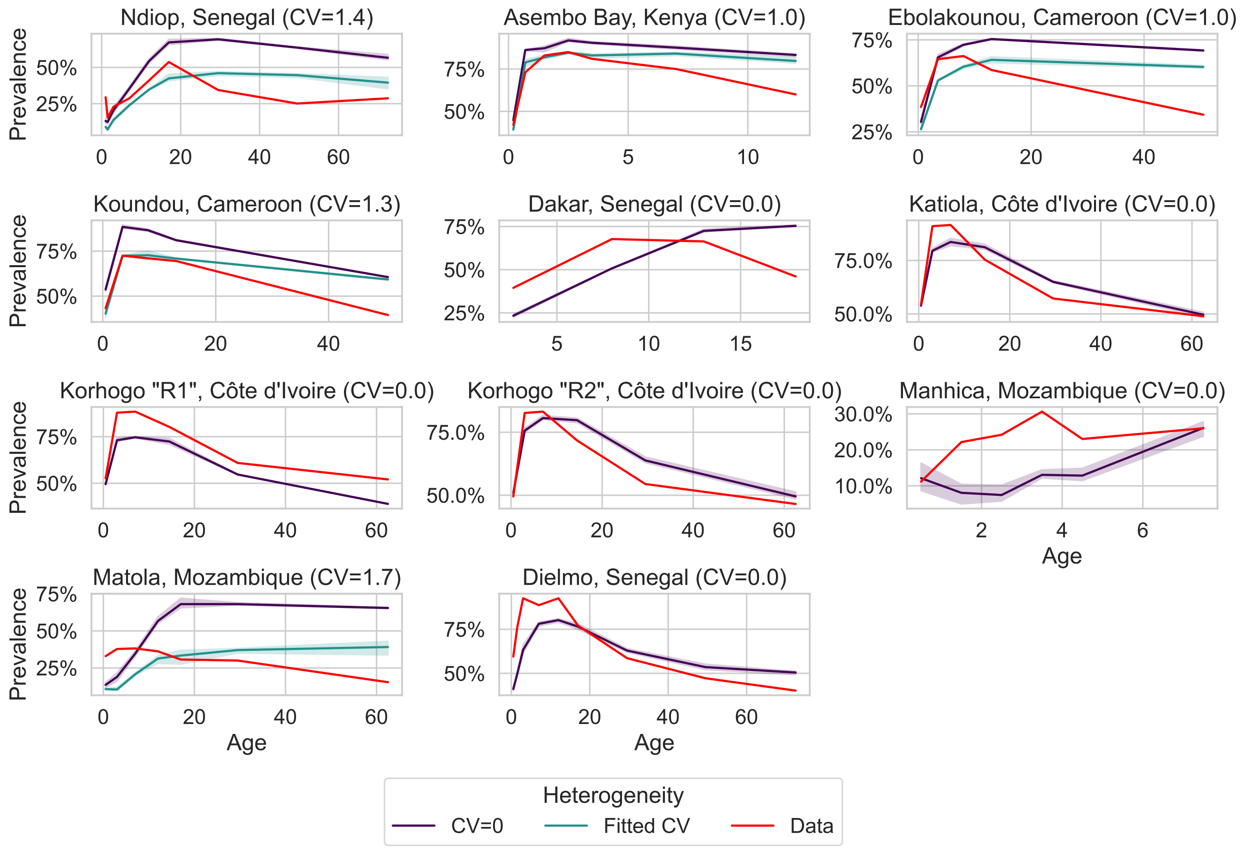


Figure S9: Age-prevalence curves collected by Battle et al. [1] used for the fitting of OpenMalaria. Heterogeneity was fitted through the coefficient of variation (CV) such that the distance between data and the curve generated by OpenMalaria was minimized as measured by sum of squares. The fitted CV value is given in brackets behind the scenario names. The improvement of the fit to the data with heterogeneity compared to no heterogeneity is given in Table S1.
